# Transformers Enhance the Predictive Power of Network Medicine

**DOI:** 10.1101/2025.01.27.25321204

**Authors:** Jonah Spector, Andrés Aldana, Michael Sebek, Joseph Ehlert, Christian DeFrondeville, Susan Dina Ghiassian, Albert-László Barabási

## Abstract

**Background:** Self-attention mechanisms and token embeddings behind transformers allow the extraction of complex patterns from large datasets, and enhance the predictive power over traditional machine learning models. Yet, being trained to make predictions about individual cells or genes, it is not clear if transformers can learn the inherent interaction patterns between genes, ultimately responsible for their mechanism of action. We use Geneformer, pretrained on single-cell transcriptomes, to ask if transformers can implicitly capture molecular dependencies, including protein-protein interactions (PPIs), allowing us to explore the use of transformers to improve network medicine tasks such as disease gene identification and drug repurposing.

**Methods:** We extracted the cosine similarity of gene embeddings and the attention weights contained in Geneformer, allowing us to test if these weights capture experimentally validated protein interactions. Using dilated cardiomyopathy as a case study, we evaluated the effectiveness of the resulting weighted networks in disease module detection and drug repurposing.

**Results:** We found that Geneformer displays awareness of experimentally documented Protein-Protein Interactions, exhibiting higher cosine similarity and attention weights for gene pairs with physical interactions. Weighting PPI networks with the cosine similarity and attention weights improved the detection of disease-associated genes and the accuracy of drug repurposing predictions for dilated cardiomyopathy, surpassing the accuracy of unweighted networks. Finally, we find that combining attention weights and cosine similarities with ranking methods enhances drug candidate prioritization for drug repurposing.

**Conclusions:** We find that transformers, by implicitly learning the interactions between genes, offer a promising pathway for advancing medicine and drug discovery when integrated with the graph theoretic algorithms used in network medicine.

## Introduction and Background

Transformers [1], a class of deep learning models designed for natural language processing (NLP), are revolutionizing biological research by capturing complex patterns in data through self-attention mechanisms. The transfer learning approach behind transformers has been applied to improve protein structure prediction [2], genetic sequence analysis [3, 4], and molecular interaction prediction [5], and to extract knowledge from large datasets [3, 6, 7].

In NLP, models are given inputs in the form of tokens, which are vector representations of the words in the sentence. In Geneformer, a transcriptome-based transformer pretrained with 30 million single-cell transcriptomes, genes are the individual tokens. While in language the order of the tokens is predefined by the grammar, Geneformer uses Rank-Value Encoding (RVE) to order genes by normalized expression defining the ‘grammar’ of the cell [7]. The pretraining gives Geneformer a versatile context awareness that can be harnessed through posterior task-specific training phases (fine-tuning).

Machine learning models are typically trained to predict individual tokens or classify groups of tokens, ignoring the inherent interactions between them. In biology, however, the interactions between genes are responsible for the mechanism of action of the individual genes and proteins. In this study, we ask to what degree transformers capture the known molecular dependencies between proteins, allowing us to explore the utility of transformers to enhance the tool set of network medicine. Indeed, the activity pattern of a gene is the result of multiple network-based dependencies, that are mapped and studied by network medicine. By conceptualizing biological processes as networks of genes, proteins, and RNAs [8, 9], network medicine offers a series of graph theoretic tools to predict drug repurposing opportunities or drug responses [10, 11, 12, 13]. Here we show that Geneformer, a six-layer transformer trained with single-cell transcriptomes, develops an awareness of experimentally documented Protein-Protein Interactions (PPI). This awareness allows us to adapt the transformer to perform standard network medicine tasks, finding that it can improve the performance of disease gene discovery and drug repurposing. Overall, our research reflects a convergence of network medicine and deep learning, offering a path towards a better understanding of complex diseases by providing predictive models of biological interactions and disease mechanisms that can help develop more effective treatments.

## Results

### The Human Interactome

Network medicine unveils disease mechanisms by mapping out the physical interactions between cellular components (genes, proteins, RNAs). Geneformer, trained on expression data, lacks direct information on these physical interactions. However, the experimentally observed expression pattern of each gene is determined by regulatory processes driven by the underlying physical interactions. Therefore, we hypothesize that information about physical interactions is implicitly encoded in Geneformer. To test this hypothesis, we examined whether an experimentally validated network of protein interactions can be extracted by exploiting the relationships between their corresponding internal representations within Geneformer. This ground truth interactome is represented as a network of 18,260 protein-coding genes (nodes) and 520,009 experimentally validated physical interactions (edges) [14]. By filtering for the 18,062 genes common to Geneformer’s vocabulary and the interactome, we derive a PPI network with 514,674 interactions. This network serves as the foundation for the analyses presented in this paper.

### Embeddings Predict Human Interactome

Geneformer encodes genes in 256-dimensional vectors known as embeddings [7]. These vectors are initially oriented randomly and change direction during pretraining, updated as they progress through each layer of the model (see Fig. 1a-c). After pretraining, the model develops a vocabulary of 25,424 genes, each corresponding to a 256 dimensional embedding that represents a single protein coding gene or miRNA. Cosine similarity of these vectors was previously used for *in silico* network analysis of congenital heart disease [7], prompting us to hypothesize that cosine similarity may reflect the underlying physical interactions between gene pairs (Figure 1c).

**Figure 1:**
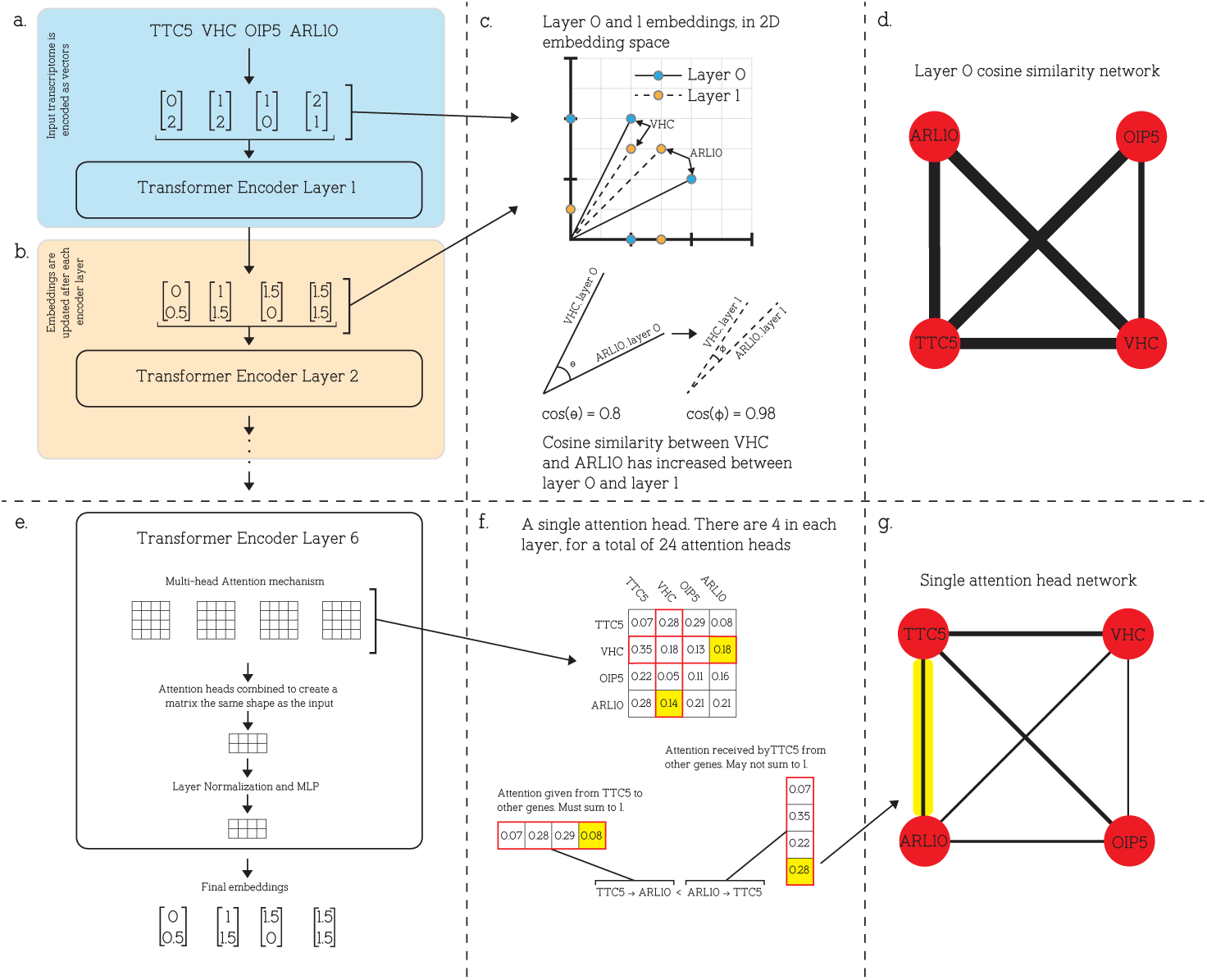
Similarity Measurements in Geneformer. **(a)** Starting with a transcriptome, genes are tokenized into vector embeddings and fed through the first transformer encoder layer. **(b)** Each layer outputs an updated set of embeddings before passing them into the next layer. **(c)** The initial embeddings (layer 0) and the layer 1 embeddings are plotted to show how the genes can move in embedding space between layers. In this case, VHC and ARL10 get closer in embedding space after layer 1. **(d)** The layer 0 embeddings are turned into a cosine similarity network, where each gene is connected to every other gene with an edge weighted by its cosine similarity. Here, ARL10 and OIP5 are orthogonal in embedding space, so no edge exists. **(e)** The final layer of the model is expanded to show the multi-head attention mechanism. All six layers have an identical mechanism, giving the model 24 total attention heads. **(f)** An attention head from the 6th layer is magnified. These attention weights are another way to compare gene similarities, as each gene attends to every other gene in the sequence. Since the matrix is asymmetric, the attention to TTC5 from ARL10 is different from the attention to ARL10 from TTC5. **(g)** The layer 6 attention weights are turned into a weighted directed network. This is an example of maximum aggregation, where we take the larger of *a_ij_, a_ji_* as the edge weight between *i* and *j*.

We run 10,000 healthy single-cell transcriptomes from the 30M corpus through the pretrained model. For each sample, we extract the embeddings from the second to last layer and calculate the cosine similarity matrix *C* that records the maximum cosine similarity between each gene pair across the 10,000 samples (Methods). If the embeddings are informed by the physical interactions between the genes, we would expect that gene pairs with experimentally validated binding interactions would have larger cosine similarity. In line with this hypothesis, we find that the mean value 0.219 *±* 0.001 of the cosine similarities (CSs) corresponding to experimentally validated PPIs is significantly different from the background (nonexistant interactions) with the mean value 0.179 *±* 0.001, a difference whose statistical significance is verified by the Kolmogorov-Smirnov test (KS-test, *p* = 0) (Figure 2a).

**Figure 2:**
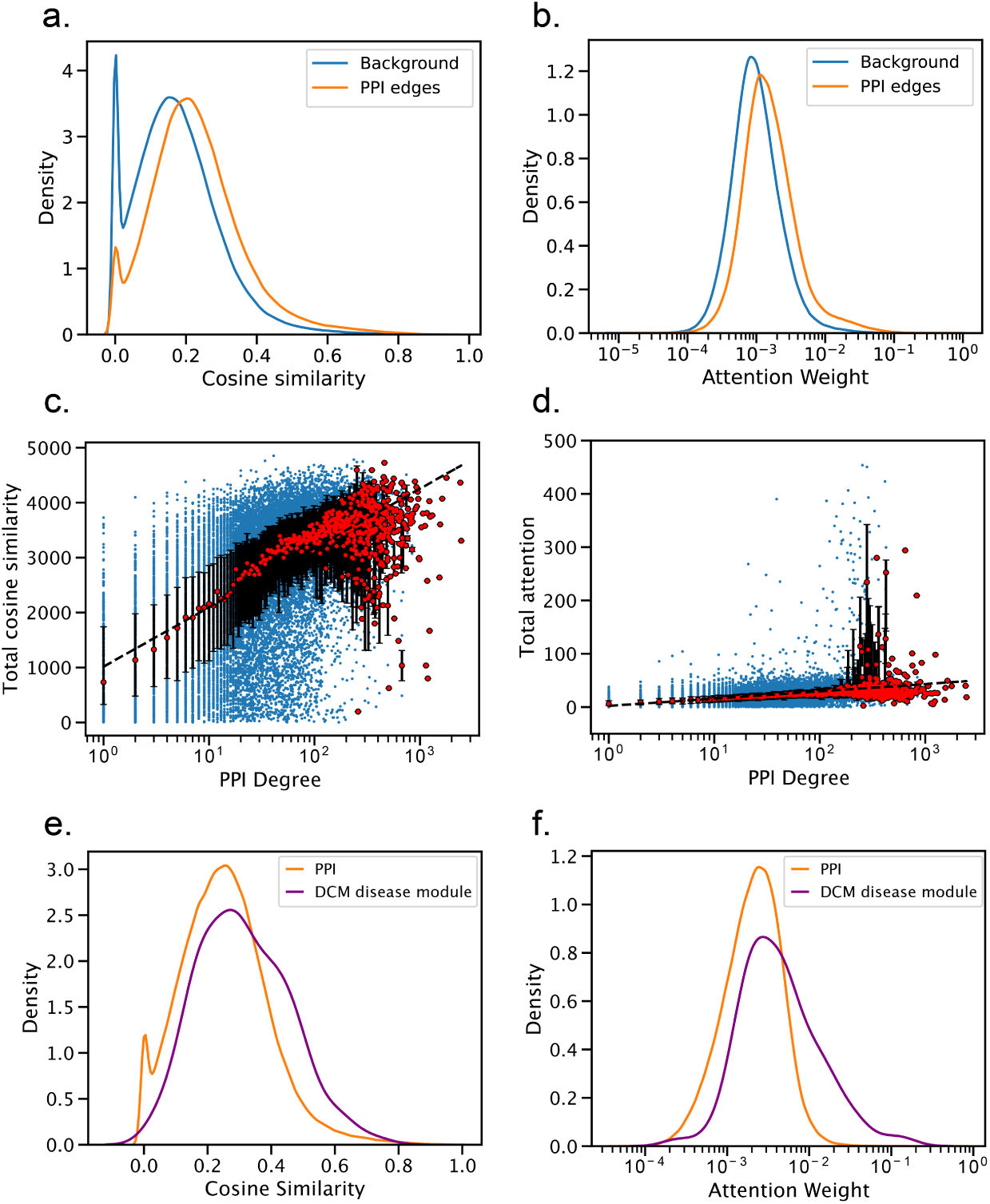
Geneformer Prioritizes PPI Edges. **(a)** Using 10,000 cells from Genecorpus, we plot distributions of cosine similarities for the PPI and the fully connected background, sobserving a higher median weight in the PPI edges. **(b)** We plot the same distributions using attention weights and again find a higher median weight in the PPI edges. **(c)** Summing across rows of the cosine similarity matrix *C*, the total cosine similarity for a gene is plotted against its degree. We find that the total cosine similarity increases with the degree, and when modeled as a logistic regression, *ln*(*k*) explains 27% of the variation in *C*, the total cosine similarity. The red points with black error bars are the mean and standard deviation across all genes with a given degree *k*, and the black dashed line is the resulting fit from the logistic regression. The blue points represent individual genes. **(d)** Summing across rows of the attention matrix *A*, the total attention for a gene is plotted against its degree. We find that the total attention increases with the degree, and when modeled as a logistic regression, *ln*(*k*) explains 9% of the variation in *A*, the total attention. **(e)** Using 10,000 dilated cardiomyopathy (DCM) cardiomyocytes, we create a new cosine similarity matrix *C* and observe that cosine similarities within the largest connected component (LCC) of DCM disease genes in the PPI (also known as the disease module) have a higher median weight than other PPI edges. **(f)** We generate a new attention matrix *A* from these same 10,000 DCM cells, observing again that the DCM disease module edges have a higher median weight than other PPI edges.

### Attention Weights Also Predict Interactions

“Attention is All You Need” stated the publication that introduced the transformer architecture [1]. Indeed, the attention mechanism helps the transformer dynamically focus on different parts of the input data by weighting the significance of each input element differently, determining how much “attention” each part needs.

Each of Geneformer’s six layers contains four attention heads (Figure 1e-f) [7]. Each head is a square matrix whose dimensions match the length of the input transcriptome, capturing the “attention” each gene assigns to every other gene within that transcriptome. We created an aggregated attention matrix *A* from the same 10,000 single-cell samples used to generate the cosine similarity matrix *C* (Methods). We find that the PPI edges have a higher mean attention weight (AW) (2.25 *±* 0.04 *×* 10*^−^*^3^) than the background (1.62 *±* 0.02 *×* 10*^−^*^3^), whose significance is again verified by a KS-test (*p* = 0).

### Degree Dependence of Cosine Similarity and Attention Weights

Given the central role the node degree *k* (number of physical interactions with other nodes) has in network medicine, we explored the relationship between the total attention and cosine similarity associated with a gene and the degree *k* of that gene in the network. By summing along the rows of the cosine similarity matrix *C*, we calculated the total cosine similarity *C_tot_* associated to each gene. Similarly, we calculated the gene’s total attention *A_tot_*. By fitting the logistic regression,

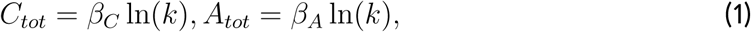

we found that the total cosine similarity and attention of each gene have a strong positive correlation with the gene’s degree *k*. *C_tot_* shows a dependence on ln(*k*) with an *R*^2^ of 0.27, while *A_tot_* has an *R*^2^ of 0.09 with ln(*k*), indicating that the natural log of the degree explains 27% of the variance in *C_tot_*and 9% in *A_tot_* (Figure 2d).

Taken together, we find that both cosine similarities and the attention weights show an awareness of the underlying physical interactions within the cell. While the resulting predictive power for protein interactions is relatively modest (Figure S4), the higher cosine similarity and attention weights associated with these interactions prompt us to ask if the transformer technique may be able to contribute to the traditional network medicine tasks, like disease module identification and drug repurposing, questions that we explore next.

### Pretrained Model Pays Attention to Disease Modules

A foundational result in network medicine is that the disease genes associated with a specific disease or phenotype are not randomly distributed in the interactome, but tend to be located in a specific network neighborhood, forming a connected subgraph known as the disease module [15, 16]. We hypothesize that attention weights and embeddings, by revealing relationships between disease genes, highlight the specific network neighborhood within the interactome, helping us identify the disease module.

To test this hypothesis, we start from a gene-disease association (GDA) list consisting of 110 disease genes associated with dilated cardiomyopathy [10]. Of the 110 proteins, 74 form a connected subgraph in the PPI, corresponding to a disease module connected by 174 physical interactions. We take expression data from 10,000 dilated cardiomyopathy (DCM) cardiomyocytes [17] and run them through the model to create a new aggregated cosine similarity matrix *C^′^*. We categorize the obtained CSs between gene pairs corresponding to the 174 edges within the disease module and those corresponding to all physical interactions in the PPI. We find that the CSs within the disease module have a mean weight of 0.313 *±* 0.002, slightly larger than 0.253 *±* 0.001, observed for all PPI interactions (Figure 2e), a difference significant by KS-test (*p* = 6 *×* 10*^−^*^5^). Similarly, we find that the attention weights within the disease module have a mean weight of 9.29 *±* 0.19 *×* 10*^−^*^3^, which is 3 times larger than the mean weight of all PPI interactions (2.78 *±* 0.003 *×* 10*^−^*^3^, Figure 2f, statistically significant by the KS-test, *p* = 2 *×* 10*^−^*^14^).

### Improving Disease Module Detection

Next, we ask if we can use Geneformer to improve disease gene predictions [18], a central task for network medicine, particularly pertinent for under-studied and rare diseases with a limited number of associated genes. Current network medicine algorithms for identifying the disease module aim to construct a connected component of disease genes in the network, relying on the unweighted protein-protein interactome (PPI) [19]. While a weighted interactome offers myriad potential improvements in network medicine [8, 10, 19], weighting techniques can introduce batch effects [20, 21], methodological differences [22], or low confidence edges [23].

Here, we hypothesize that an interactome weighted with either Geneformer AWs or CSs will allow us to improve network medicine tasks such as disease module discovery. The leading algorithm for disease module detection, random walk with restart (RWR) [24, 25, 26], begins with a set of known disease genes acting as seeds and then ranks new candidate genes in the interactome by calculating the steady-state probability that a walker, starting from any seed gene, will reach each candidate gene. The probability that the walker reaches a candidate gene defines the probability that the gene should be a part of the disease module.

We applied the aggregated Geneformer AWs from the pretrained model as weights to the PPI edges. To be specific, for each link (*i, j*) in the PPI, the interaction strength is defined as *W_ij_* = max(*A_ij_, A_ji_*), where *A_ij_*represents the aggregated attention weight between genes *i* and *j*, derived from the transcriptomes of 10,000 cardiomyocytes from dilated cardiomyopathy (DCM) samples. We repeated this process using AWs from the Geneformer model fine-tuned on cardiomyopathy. Finally, we repeat the entire process using cosine similarities (CSs), for a total of 5 networks (unweighted, pretrained embedding weighted, pretrained attention weighted, fine-tuned embedding weighted, and fine-tuned attention weighted.) We divided the set of 110 known cardiomyopathy genes into seed (80%) and recovery (20%) sets. The seed set served as the starting point for RWR, while the recovery set is used to measure the predicted rank of the known disease genes.

We find that all 4 weighted networks perform at least as well as the unweighted network, and the attention weighted networks offer a notable improvement in disease module detection. Both the pretrained and fine-tuned attention weighted networks generate an AUROC of 0.76 *±* 0.02, which is within the standard error in the mean of the unweighted network (AUROC = 0.74 *±* 0.02, Figure 3a). For the purposes of disease module detection though we are mainly interested in the top candidates. Therefore, we evaluated the precision and cumulative true positives in the top 100 candidates (Figure 3c,d), since these metrics can represent the efficacy of the algorithm more accurately than the AUROC. We find that in the top 100 candidates, the attention weighted networks record nearly twice the cumulative true positives compared to the unweighted network(Figure 3d). The fine-tuned AWs in particular generate an initial precision of nearly 60%, a notable improvement on the unweighted network, which records an inital precision of 0 (Figure 3b,c).

**Figure 3:**
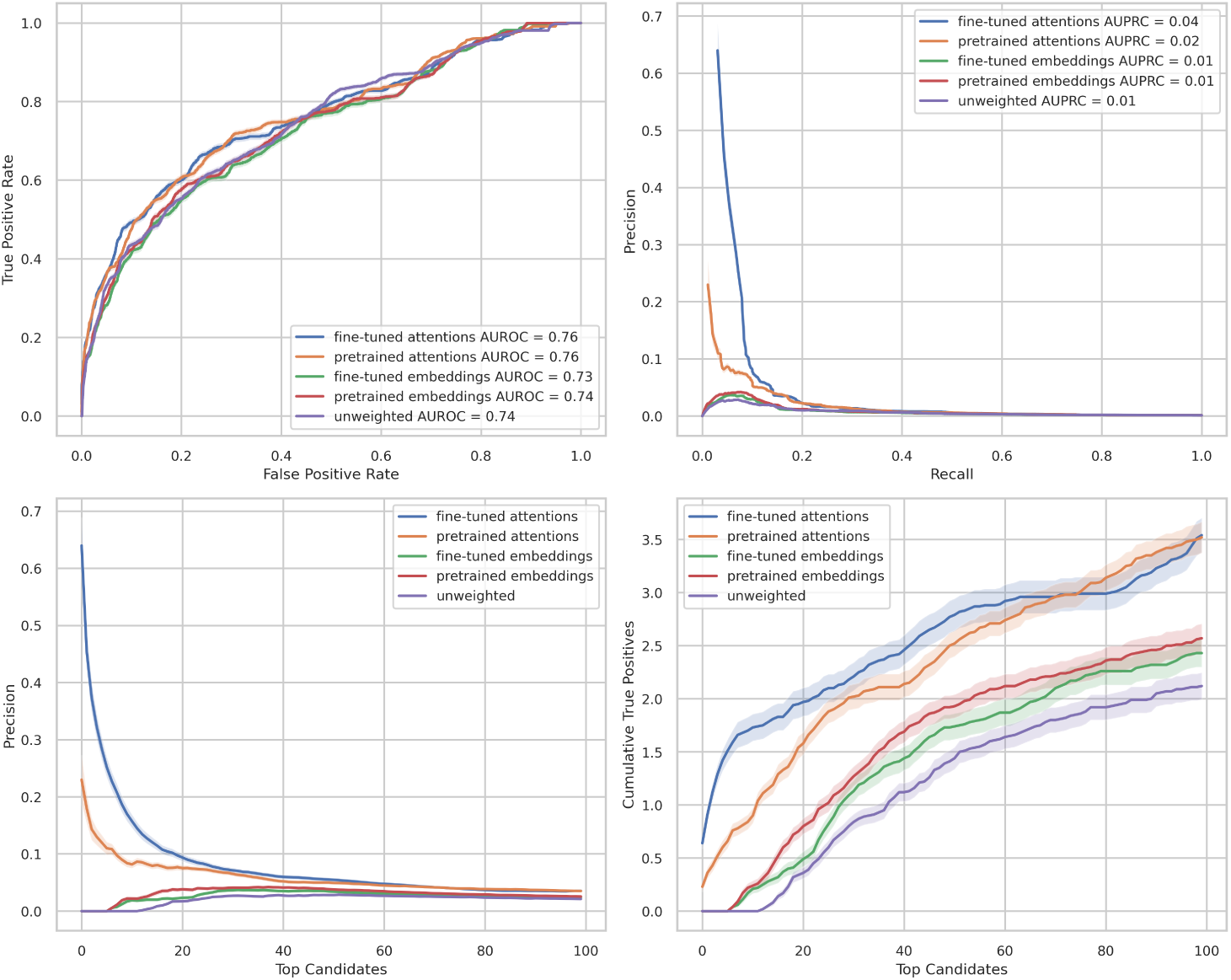
Geneformer Attention Weights Improve Capture-Recapture Analysis for Disease Module Discovery. **(a)** ROC curves are plotted for 4 different Geneformer weighted networks and a control unweighted network, comparing their recovery of removed disease module genes. The shaded ribbon indicates standard error in the mean of 100 trials with different seed genes. **b)** The Precision-Recall curve is plotted for each network. The attention weighted networks have considerably higher initial precision than other networks, prompting us to look more closely at this region. **(c)** The precision is plotted for the top 100 candidates. The fine-tuned attention weighted network has higher precision for the top 100 candidates than any other network, followed by the pretrained attention weighted network. **(d)** The cumulative true positives are plotted for the top 50 candidates. The attention weighted networks record the most true positives (between 3 and 4 on average), but the cosine similarity weighted networks also outperform the unweighted network.

### Attention Weights Improve Drug Repurposing

Drug repurposing, the process of identifying new therapeutic uses for FDA-approved drugs, has become increasingly important due to its potential to reduce the time and cost associated with drug development. While some approaches leverage computational methods and extensive biological databases to predict drug-disease associations [27, 28, 29], network medicine based repurposing, using either graph theory tools [11, 13], graph neural networks [30], or a combination of these two methods [10], was able to identify promising experimentally tested drug candidates, along with the specific disease mechanism responsible for the pathogenesis [31].

We evaluated whether the AWs and CSs between Geneformer embeddings could improve drug repurposing accuracy. As a baseline, we used the dilated cardiomyopathy (DCM) finetuned Geneformer model [7]. We extracted the AWs and CSs from the model and used them to assign weights to the PPI network. Subsequently, we calculated the network-based proximity scores [32] between the DCM disease genes and the targets of 618 drugs, including 171 DCM-related (Positive) and 447 unrelated (Negative) drugs extracted from DrugBank (see Methods) [33]. We ranked the drug list according to each drug’s proximity score and tracked the number of positive and negative drugs predicted by the DCM-Geneformer weighted networks.

We repeated this procedure by extracting the AWs and CSs values from all six transformer layers of Geneformer. As a control, we compared the results from these weighted networks with those from the unweighted PPI network. We find that the PPI network weighted with AWs from layer 5 and CSs from layer 0 enhanced the overall prediction capabilities for identifying DCM-related prescribed drugs, achieving an AUROC of 0.72 for AWs and 0.71 for CSs. This performance considerably surpassed the predictive power of the unweighted PPI network (AUROC = 0.60), as well the predictive power extracted from the other layers (Figures 4a,d and g).

**Figure 4:**
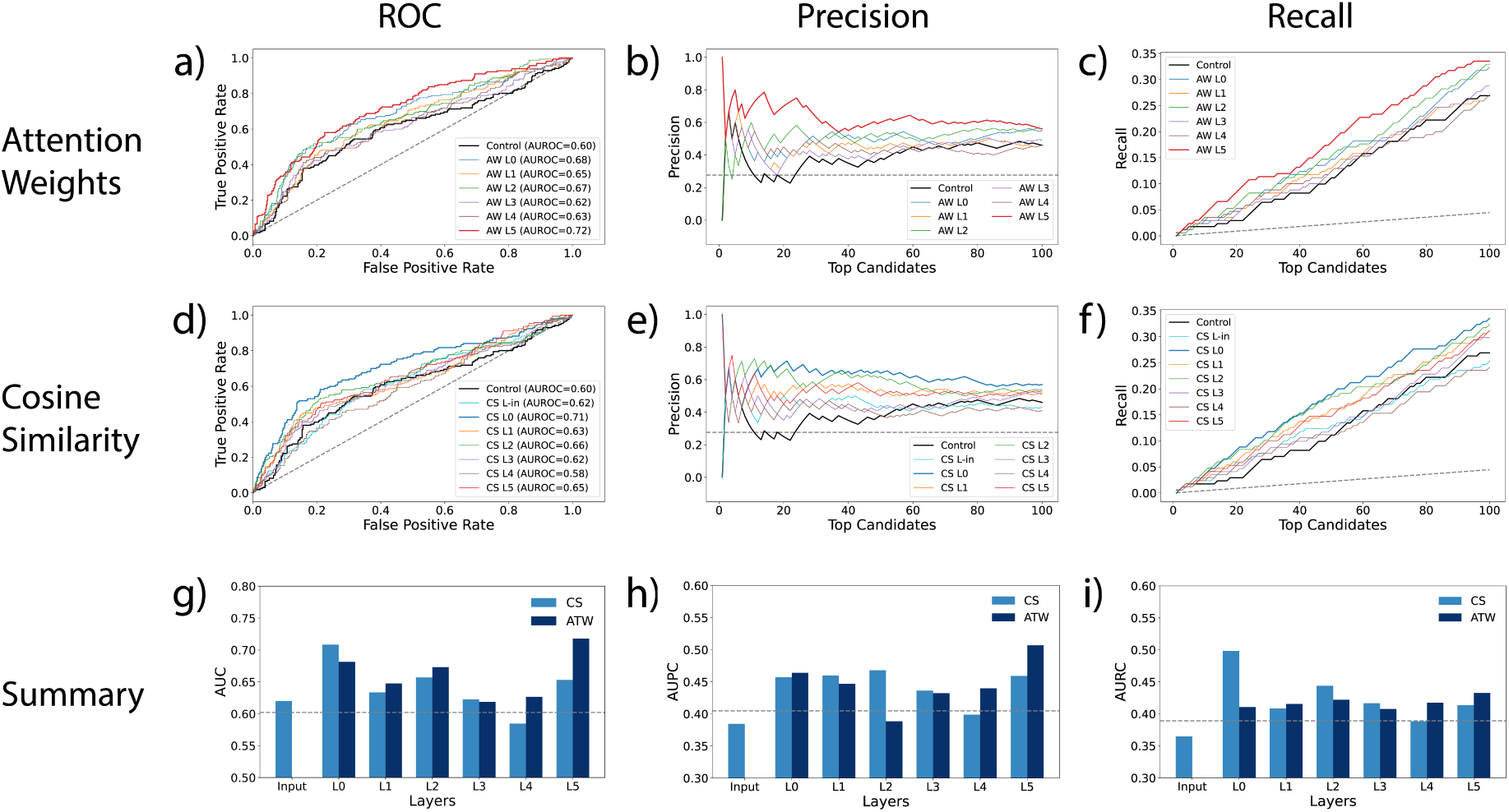
Layer-Wise Comparison of Geneformer Attention Weights and Cosine Similarity for Drug Repurposing in Dilated Cardiomyopathy. **a, d)** ROC curves comparing the drug repurposing performance using attention weights (AWs) and cosine similarities (CSs) from six Geneformer layers. AWs from layer 5 (AUROC = 0.72) and CSs from layer 0 (AUROC = 0.71) show the best predictive capacity in their respective sets, with both outperforming the unweighted control. **b, e)** Precision curves for the top 100 drug candidates. AWs from layer 5 (AUPC = 0.50) and CSs from layers 0 (AUPC = 0.45), 1 (0.45) and 2 (0.46) maintain the highest precision across most top candidates, indicating better predictive power compared to other layers and the control. **c, f)** Recall curves for the top 100 drug candidates using AWs and CSs from different layers. AWs from layer 5 (AURC = 0.43) and CSs from layer 0 (AURC = 0.50) achieve the highest recall, identifying more known cardiomyopathy drugs in the top candidates compared to other layers and the control network. **g-i)** AUROC, AUPC and AURC values comparing AWs and CSs across Geneformer layers. The gray dashed line represents the respective values derived from the unweighted control network (AUROC = 0.60, AUPC = 0.40, AURC = 0.39). In all cases, the results represent the average of ten independent runs, with a standard deviation below 0.02.

The aim of drug repurposing is to prioritize the available drugs, so that experimental efforts can be focused on the highest-ranked compounds. For this reason, we calculated the number of true positive drugs among the top 100 predicted drugs (precision), and the fraction of all possible positive drugs among the top 100 predicted drugs (recall). For precision, the PPI network weighted with AWs from layer 5 (AUPC = 0.43) and CSs from layer 0 (AUPC = 0.45) outperformed both the unweighted control network (AUPC = 0.40) and other layers. Similarly, for recall, the PPI network weighted with AWs from layer 5 (AURC = 0.43) and CSs from layer 0 (AURC = 0.50) also surpassed the control network (AURC = 0.39) and other layers (Figure 4b,e,h and 4c,f,i).

Since AWs from layer 5 and CSs from layer 0 demonstrated the best overall performance from each set, we further explored the relationship between their drug proximity scores. As shown in Figure 5a, the scores derived from AWs and CSs exhibit a strong positive correlation (Pearson correlation coefficient = 0.77, *R*^2^ = 0.59), indicating that both methods capture similar network-based relationships between drug targets and DCM genes. However, disagreement between the two scores suggests that each method captures different aspects of the relationships, indicating their potential complementarity for improving prediction accuracy when combined. To assess this, we applied three ranking methods: Borda Count [34], Dowdall Count [35], and CRank [30], to integrate the scores provided by AWs and CSs. These combined ranking methods yielded a marginally higher predictive performance than AW or CSs scores alone, achieving AUROC values of 0.74 for Dowdall Count and CRank (Figure 5b). Additionally, precision and recall analyses of the top 100 ranked drugs revealed that CRank is the most effective combination method, outperforming Borda and Dowdall counts, as well as the individual AW and CSs scores and the unweighted PPI network. This suggests that integrating multiple layers of information from Geneformer enhances the accuracy of drug repurposing predictions (Figure 5c,d).

**Figure 5:**
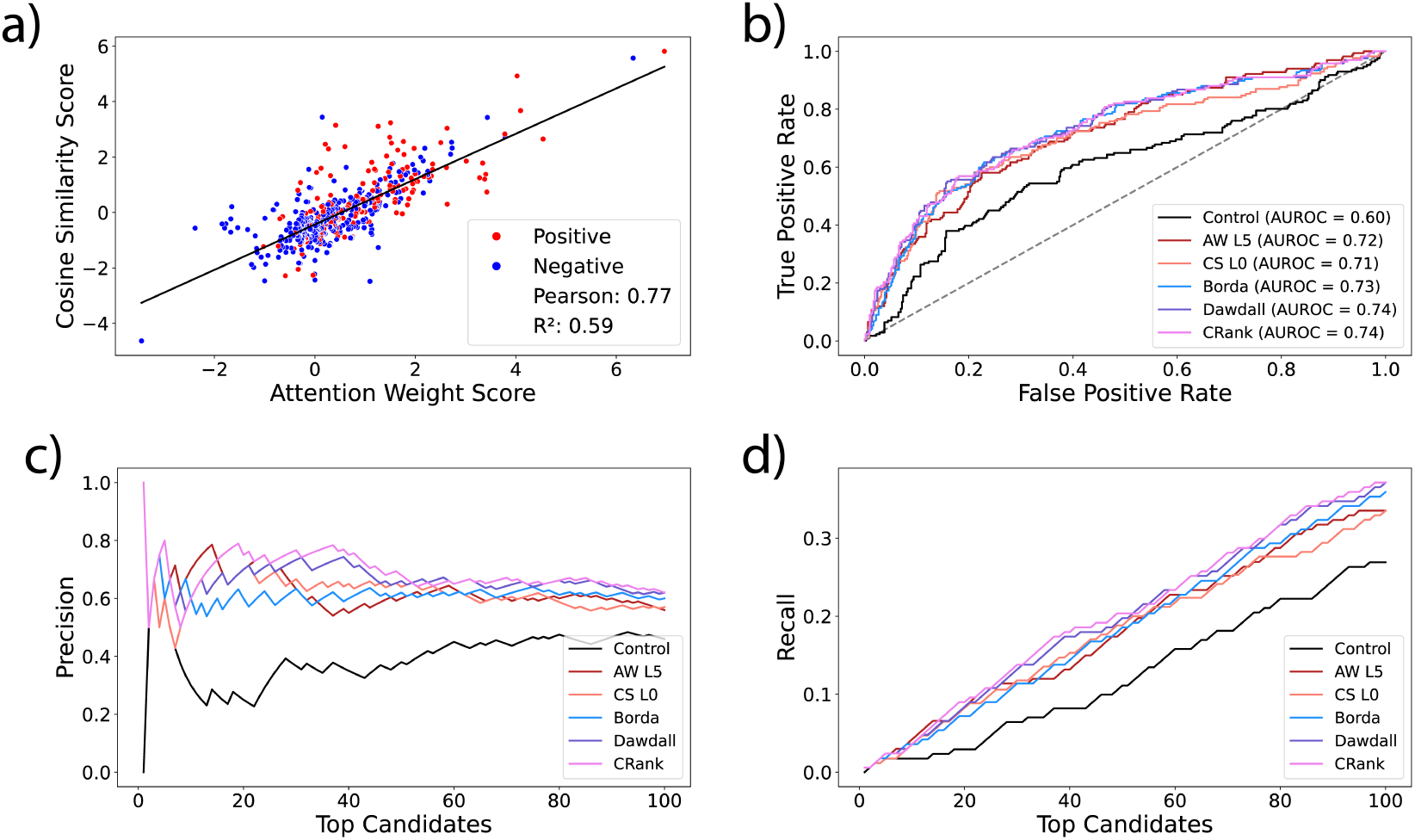
Correlation and Performance of Drug Repurposing Scores Derived from Cosine Similarity (CSs) from Layer 0 and Attention Weights (AWs) from Layer 5 Using Combined Ranking Methods. a) Correlation between drug repurposing scores derived from CSs from layer 0 and AWs from layer 5. The score is calculated as *−*1 *×* proximity(DCM, D), where DCM represents dilated cardiomyopathy disease genes, and D represents the drug targets. Blue points correspond to negative drugs, and red points correspond to positive drugs used to treat cardiomyopathy. The *R*^2^ value is 0.59. b) Receiver Operating Characteristic (ROC) curves comparing the performance of drug repurposing using individual scores from CSs from layer 0, AWs from layer 5, and combined scores generated using Borda Count, Dowdall Count, and CRank methods. The combined ranking methods show improved performance, with CRank (AUROC = 0.74) and Dowdall (AUROC = 0.74) achieving the highest AUROC values. c) Precision curves for the top 100 drug candidates ranked by proximity to cardiomyopathy genes. The combined ranking methods show improved precision over the individual CS and AW scores, with CRank maintaining higher precision across most top candidates. d) Recall curves for the top 100 drug candidates, comparing individual and combined scores. The combined scores, particularly CRank and Dowdall, achieve higher recall, identifying more known cardiomyopathy drugs in the top-ranked candidates. In all cases, the results represent the average of ten independent runs, with a standard deviation below 0.02.

## Discussion

In this study, we demonstrated that transformer-based models, specifically Geneformer, have the potential to enhance network medicine tasks, such as disease gene identification and drug repurposing. Our results illustrate how transformer embeddings and attention weights can be leveraged to capture and reinforce biological interactions within protein-protein interaction (PPI) networks. While network medicine traditionally relies on unweighted or experimentally weighted networks [36, 37, 38], we show that attention-weighted networks outperform traditional methods.

A key finding of this study is the superior performance of attention weights compared to cosine similarity in disease gene identification and marginally in drug repurposing. While both mechanisms capture relationships between gene pairs, we find that attention weights outperform cosine similarity in accuracy and applicability.

One possible explanation for this difference is that attention weights are dynamic and context-dependent, in contrast with the more static, global relationships between gene embeddings offered by cosine similarity [1]. This dynamic nature of attention weights makes them particularly well-suited for tasks like disease module detection and drug repurposing, where functional relationships between genes are context-specific.

We also find that fine-tuning Geneformer on disease-specific data, such as cardiomyopathy transcriptomes, further enhances its ability to identify disease-relevant network regions. In our analysis of the DCM disease module, the fine-tuned model’s cosine similarity values and attention weights showed greater separation between PPI and disease module edges than the pretrained model (Figure S5). This increased focus on the disease module, alongside with a down-prioritization of unrelated interactions, highlights the value of fine-tuning transformer models for specific diseases.

One of the most impactful applications of Geneformer’s attention weights and cosine similarity is in drug repurposing. Our results show that AWs from layer 5 and CSs from layer 0 provided the highest overall accuracy, outperforming both the unweighted PPI network and other layers. Moreover, integrating AWs and CSs from the Geneformer model slightly improves drug repurposing predictions for dilated cardiomyopathy (DCM). While AWs and CSs scores are strongly correlated, their differences suggest they capture complementary information, with combined ranking methods like CRank [30] further enhancing predictive power. These findings highlight the value of integrating transformer models with network medicine approaches, demonstrating that attention weights and cosine similarity together can improve the identification of therapeutic candidates for complex diseases. While Geneformer’s performance in network medicine tasks is promising, limitations remain.

Though improved, the AUROC values indicate room for further optimization, particularly in integrating additional biological data or refining the transformer model architecture. Additionally, exploring other deep learning architectures or expanding the training datasets could help improve the accuracy of disease gene identification and drug repurposing predictions. Further research is needed to determine how these transformer-based models can be applied to more diverse biological contexts, including signaling pathways or metabolic networks.

This convergence of AI and network medicine offers unprecedented opportunities for understanding complex diseases. Using transformers in network biology could significantly accelerate drug discovery efforts, reduce the cost and time of drug development, and improve patient outcomes through more targeted therapeutic interventions.

## Methods

To evaluate Geneformer’s ability to capture biological information, we compared embeddings and attention weights from a pretrained and fine-tuned model with a protein-protein interaction (PPI) network and curated disease-gene association lists (SI 1.1). We sampled 10,000 single-cell transcriptomes and calculated the aggregated cosine similarity and attention weights to assess the distribution of known PPI and disease module edges (SI 1.2). We further used the cosine similarity and attention weight values to construct weighted PPI networks (see SI 1.3). We used the weighted networks to perform disease module discovery (SI 1.3) and drug repurposing studies (see SI 1.4) in dilated cardiomyopathy.

## Data Availability

All raw datasets used in this manuscript are directly cited. The source code is available at https://github.com/Barabasi-Lab/Geneformer-NetworkMedicine/tree/main, along with instructions for recreating and replicating the analyses in the paper. The aggregated weight matrices used to create the figures are available at https://huggingface.co/datasets/jspector792/Geneformer_NetworkMedicine.

## Acknowledgments

We acknowledge Katarina Petrovich, Giulia Menichetti, and Abel Elekes for valuable discussions, and Christina Theodoris for advice related to the internal workings of Geneformer.

## Materials & correspondence

Correspondence and requests for materials should be addressed to A.L.B.

