## Supplementary Information for "Transformers Enhance the Predictive Power of Network Medicine"

### Transformers Enhance the Predictive Power of Network Medicine - Supplementary methods and results

**2 Choosing a layer** **6**

**3 Combining Attention Weights and Embeddings** **7**

**4 PPI Edge Analysis** **8**

**5 Geneformer Fine-Tuned With Disease Transcriptomes Focuses More Attention to the Disease Module** **11**

**6 Attention Weights Do Not Identify Physical Disease Gene Interactions** **13**

**7 The Non-coding Interactome** **15**

### 1 Methods

#### 1.1 Aggregating Weights from Geneformer

In order to impose Geneformer attentions weights (AWs) or the cosine similarities of its embeddings (CS) on the protein-protein interactome (PPI), we create a large database of AWs and CS from which we can sample PPI edges. To get an AW or CS between two genes $i$ and $j$, they must be co-expressed in a cell (both so that they are biologically related, and because this is the only way to generate an AW or CS between two genes). Therefore, we randomly select 10,000 transcriptomes from a dataset (either the pretraining dataset, Genecorpus 30M; or the fine-tuning training data subset of 172,000 dilated cardiomyopathy (DCM) cardiomyocytes [1]). We select 10,000 because it is the largest set of samples we can feasibly aggregate on a regular basis. We verify in Figure S1 that 10,000 samples is enough to reach a regime of diminishing returns in the number of unique genes we find in the sample set. We create an $N \times N$ matrix (we refer to this as $A$, but in all applications it is interchangable with $C$) to hold our aggregated weights, where $N$ is the total number of unique genes in the 10,000 sample set. We then run these 10,000 samples through the model (either the pretrained Geneformer, or the fine-tuned cardiomyopathy modelprovided by [2]). For each sample we extract the CS and AWs between each pair of genes in the sample. We then update $A$ using either maximum aggregation or mean aggregation.

For maximum aggregation, we simply record the largest AW between genes $i$ and $j$ in $A_{ij}$. For mean aggregation, we take the average value across all co-occurrences of $i$ and $j$. When we are aggregating AWs, we rescale each attention weight by a factor of $l/2048$, where $l$ is the length of the transcriptome. Since attention matrices are row normalized, this ensures that AWs coming from shorter transcriptomes do not dominate. After this process, we have a matrix of the gene-gene relationships. In the DCM dataset, we find an average of 17,222 genes per 10,000 samples, with a standard deviation of 20 across 10 different sets. In the Genecorpus 30M dataset, which contains more than 1 tissue and therefore more transcriptional diversity, we see a mean size of 20,767 genes and a standard deviation of 46 genes. We can then sample from this matrix to create a weighted PPI by taking edges in the PPI $(i, j)$ and weighting them with the corresponging Geneformer weight $A_{ij}$. These Geneformer weighted networks will inevitably lose some links compared to the ground-truth unweighted PPI, because not every interaction will appear in the dataset. However, using 10,000 Genecorpus samples we are able to recover 98% of the ground truth edges, and using 10,000 DCM samples we recover 89% of the ground truth edges, indicating that these 10,000 sample sets do cover a vast majority of the interactome.

##### 1.2 Analyzing Protein-Protein Interactions in Geneformer

Using an aggregated weight matrix $A$ we can now analyze the PPI. We use a previously reported protein-protein interaction (PPI) network with 18,062 nodes and 514,674 edges [3]. We divide the weights in $A$ into exclusive categories, based on their association with the PPI: (1) A large number (over 100 million) background edges, which do not appear in the PPI; (2) The 514,674 PPI edges; (3) The 11,926 non-physical interactions between any of the 110 dilated cardiomyopathy disease genes in the network. These genes are known from previously reported disease-gene association lists to be associated with DCM [4] and a subset of them create a connected component in the PPI. The edges of this connected component (the disease module) comprise the final set of interactions; (4) the 174 edges of the cardiomyopathy disease module. To create set (3), we take the fully connected set of disease genes (12,100 edges) and subtract the physical edges (174 edges) leaving only the non-physical edges.

#### 1.3 Disease Module Detection

We selected dilated cardiomyopathy (DCM) for our capture-recapture analysis since Theodoris et al. provide a large dataset single-cell transcriptomes. After aggregating (using maximum aggregation) AWs and CS from 10,000 cells from this dataset, we have a PPI with 16,454 nodes and 468,034 edges, approximately 91% of the original PPI. The cardiomyopathy disease module in this PPI retains all 174 of its edges, and 107 of the 110 cardimyopathy disease genes appear in both networks. We then create two weighted networks, one from the cosine similarity matrix $C$, and one from the attention weight matrix $A$. We do this by taking each edge $(i, j)$ in the PPI and weighting it with $A_{ij}$ (or $C_{ij}$ for the cosine similarity network). Because we aggregate across the diagonal in $A$, it is a symmetrical matrix even though the individual attention heads are not, leading to an undirected weighting of the PPI.

We repeat this process for both the pretrained model and the fine-tuned cardiomyopathy model, to create a total of four weighted PPIs (Pretrained AW weighted, fine-tuned AW weighted, pretrained CS weighted, and fine-tuned CS weighted), plus one unweighted. These networks are based on the same 10,000 samples, and therefore have the same underlying topology (removing the weights would make them identical).

Starting with the cardiomyopathy disease module, we randomly select 80% of the disease genes to keep. We then run a random walk with restart (RWR) on each weighted network, where at every time step the walker has a 40% chance of restarting to a randomly selected seed gene. The non-seed genes are ranked by their probability of visitation, where those nodes with the highest probability are considered the most likely to be disease genes. We set a threshold $k$ for the number of genes we consider positives, and create a ROC curve by varying $k$ from 0 to $N$, where $N$ is the number of nodes in the network (minus the seeds). We run the RWR using networkx's PageRank [5] algorithm, which is probabilistic. Therefore, we repeat this process 20 times for each network and calculate the mean and standard error. We find in main Figure 3 that the fine-tuned attention weights are the only ones that substantially improve the results of the RWR.

##### 1.4 Drug Repurposing

Using the same weighting process described above, we create several weighted Geneformer networks using weights aggregated from the fine-tuned model. In contrast to the disease module analysis, where we took weights only from layer 4, here we test all 6 layers of the model for both attention weights and embeddings, as well as the input embeddings. We extracted a set of 44 curated DCM disease genes from DisGeNet [6] with a Disease-Gene score greater or equal than 0.3. On each of the 13 weighted networks, we look at the proximity of the set $T$ of DCM disease genes to various sets $S$ of drug targets.

We extracted drug targets from DrugBank [7] for 618 drugs of which 171 drugs are currently approved for treating DCM-related conditions (Positives) and 447 drugs not used for DCM (Negatives). The Positive drugs included 7 for dilated cardiomyopathy, 1 for ischemic heart disease, 121 for hypertension, 66 for heart failure, and 20 for arrhythmia. The negative drugs were selected as those that were neither approved nor under investigation for DCM treatment and whose targets do not overlap with the targets of the 171 positives.

For each set $S$ of drug targets, we used the python library NetMedPy [8] to calculate the proximity z-score [9] between $S$ and $T$, the DCM disease genes. The proximity is a measurement of the average shortest path in the network from the genes in $S$ to the genes in $T$, which means the z-score tells us how many standard deviations closer or farther away from the disease module a given set of drug targets are. A score of $-3$ for a given set of targets indicates that those drug targets are 3 standard deviations closer to the disease module that we would expect randomly. We therefore rank the drugs in ascending order according to their proximity scores. As we did for disease module detection, we create a ROC curve by setting a threshold value $k$ for the number of drug we consider as positives from this ranked list, and then varying $k$ from 0 to 618. The results represented for drug repurposing represent the average of ten independent runs, with a standard deviation below 0.02.

### 2 Choosing a layer

Geneformer has 6 layers, each with its own set of embeddings and 4 attentions heads. The distribution of AWs and CS can vary considerably from layer to layer, so we choose one layer at a time to aggregate weights from. Initially, on the advice of Theodoris et al., we chose the 5th layer of the model (hereafter referred to by its layer index, layer 4), on the basis that it should incorporate context from the previous 4 layers, but not have task dependent features that might impact the weights from the final layer. For disease module detection, this turns out to be a prescient choice: as shown in Figure S2a,b, layer 4 AWs generates the best results in recapturing disease genes, although layer 0 and 1 AWs trail closely behind. For drug repurposing however, this assumption proved insufficient. Layer 5, assumed to be task dependent, produces AWs with the best results in drug repurposing (Figure S2c,d), and on the embeddings side we find the best performance in the layer 0 embeddings (which have no prior layers to incorporate context from). It seems that although Geneformer weighted networks generally outperform unweighted ones, the best choice of a layer is task dependent, and the precise differences in the networks created from the different layers merit future investigation.

### 3 Combining Attention Weights and Embeddings

As shown in main Figure 3 and Figure 4g,h,i of the main, AWs most often provide the best result when weighting a PPI for network medicine tasks. As Figure 4i in particular shows though, this cannot be taken for granted, embeddings can provide comparable or even superior predictive power. We also observe in Figure S3a that CS and AWs are generally not correlated (pearson correlation $= 0.16$). We therefore hypothesize that the two sets of weights are encoding different information, and combining them could yield additional predictive power. Specifically, we take the ranked list of candidate genes or drugs generated with the attention weights, and the corresponding ranked list generated with the embeddings, and for each gene or drug combine its attention rank $R_a$ with its embedding rank $R_e$. We do this using the average rank $R_A$, the Borda rank $R_B$, the Dowdall rank $R_D$, and the C rank

$R_C$.

$$R_A = \frac{R_a}{2} + \frac{R_e}{2} \quad (1)$$

$$R_B = 2N - R_a - R_e \quad (2)$$

$$R_D = \frac{1}{R_a} + \frac{1}{R_e} \quad (3)$$

$$R_C = \frac{1}{R_a^p} + \frac{1}{R_e^p} \quad (4)$$

Where in the Borda rank $N$ denotes the number of possible ranks (so a larger $R_B$ is better) and in the C rank $p$ is set to 4. In general, these methods do not extract additional predictive power compared to AWs or CS alone. In predicting whether an edge is part of the PPI or the background, we see a 1% improvement over AWs alone (Figure S3b), in drug repurposing a 2% improvement (Figure S3e,f), and in disease module detection, no improvement (Figure S3c,d).

### 4 PPI Edge Analysis

To further probe the extent to which the Geneformer weights are informed by physical interactions, we use AWs and CS aggregated from Genecorpus 30M and run through the pretrained model to examine a degree-preserving randomization of the PPI. This randomization keeps the same number of interactions for each gene, while randomizing the specific genes that are a part of the interaction. Effectively, we are creating a network that is topologically the same as the PPI, while eliminating important gene-specific biological context. Surprisingly, we find that this randomization generates a distribution of cosine similarities that is nearly indistinguishable from the real PPI. While a KS-test only tells us that the distribution of PPI weights is different from the randomized weights, the earth mover's distance (EMD) [10] sheds more light on the comparison.

The EMD, ubiquitous in machine learning [11, 12, 13], takes the distance between the distributions into account. Specifically, it measures the minimum cost to move one distribution on to the other. If two distributions move further apart, the EMD will be higher. Here, we observe that the EMD from the PPI to the degree-preserving randomization is only 0.01, compared to an EMD of 0.055 between the PPI and the controls. Because the EMD between the PPI and its degree-preserving randomization is one fifth the EMD between the PPI and the background, we can say that the PPI is roughly five times more distant to the background than the randomization.

We also test a non-degree-preserving randomization, which keeps the same number of total interactions as the PPI but randomly distributes them across all the genes, thus destroying any biological information in the network. We find that the PPI edges have a higher median weight ($1.42 \times 10^{-3}$) than the background and the non-degree-preserving randomization of the PPI ($0.94 \times 10^{-3}$ and $0.99 \times 10^{-3}$ respectively). The degree-preserving randomization has a median weight of $1.37 \times 10^{-3}$, much closer to the PPI. These comparisons are also reflected in the EMDs, where we see that the PPI is 4.5 times more similar to the degree-preserving randomization (EMD = $0.30 \times 10^{-3}$), than the background ($1.41 \times 10^{-3}$) and the non-degree-preserving randomization ($1.34 \times 10^{-3}$).

While the EMD between the PPI and the degree-preserving randomization is only 0.01, the EMD between a PPI and a non-degree-preserving randomization is 0.046, much closer to the

EMD between the PPI and the background (Figure S4a).

Because the PPI has a heavy tailed degree distribution [14], one of the effects of a non-degree-preserving randomization is to dramatically deprioritize the hubs of the network. We therefore hypothesize that the similarity between two genes is dependent upon the degrees of both genes in the PPI. This would mean that a degree preserving randomization would preserve the hubs' relative importance in the network, while a non-degree-preserving randomization would eliminate many high similarities coming from the hubs.

In the main text, we show that the total weight of a gene in the fully connected Geneformer weight matrix depends significantly on its degree in the PPI, whether the weights are cosine similarities or attention weights (main Figure 2). This supports the idea that the degree of a node in the PPI is encoded in Geneformer's internal representation.

Having found that the cosine similarities of gene embeddings prioritize PPI edges, and are predicted well by PPI degree, we now ask to what extent the cosine similarities predict the edges of the PPI. To accomplish this, we rank each pair of genes by their cosine similarity, from most similar to least similar. We then count how many physical interactions (PPI edges) appear in the top K interactions. In the top 100 cosine similarities, we recover no PPI edges, although the expected value from random selection is only 0.16. In the top 1,000 cosine similarities, we recover 6 PPI edges, almost 4 times the expected value of 1.6. Overall, we obtain an AUROC of 0.63, indicating that the gene embeddings can detect the PPI edges among the much larger fully connected background (Figure S4d).

Finally, we rank the gene pairs by their AWs, and measure how quickly we recover physical interactions using this ranked list. We find that AWs recover physical interactions faster than random expectation, and notably, faster than ranked cosine similarities. If we select edges by highest AW, we get 6 hits in the top 100 weights, compared to 0 hits from the cosine similarity list. We get 61 hits in the top 1,000, compared to only 6 hits from the cosine similarities. The AUROC is 0.65, slightly higher than what we obtain using the cosine similarities (Figure S4c).

#### 5 Geneformer Fine-Tuned With Disease Transcriptomes Focuses More Attention to the Disease Module

Geneformer also contains a fine-tuned model (GF-Cardio) trained on 93,589 transcriptomes collected from patients diagnosed as either cardiomyopathy dilated, cardiomyopathy hypertrophic, or healthy, reportedly boosting accuracy when predicting cardiomyopathy disease states [2]. We hypothesize that fine-tuning may prompt GF-Cardio to pay more attention to the network neighborhood containing the dilated cardiomyopathy disease module when it encounters DCM transcriptomes. To test this hypothesis, we repeat the above analysis with GF-Cardio, finding (Figure S5c) that while the disease module AWs shift up (from a mean of $9.29 \pm 0.19 \times 10^{-3}$ to $13.33 \pm 0.37 \times 10^{-3}$) the PPI attention weights shift downwards (from a mean of $2.78 \pm 0.003 \times 10^{-3}$ to $1.82 \pm 0.003 \times 10^{-3}$), creating greater distinction between the PPI and the disease module attention weights. We further observe that the earth mover's distance (EMD) between the PPI AWs and the disease module AWs in the pretrained model (0.007) is smaller than the EMD between the PPI AWs and the disease module AWs in the fine-tuned model (0.012) indicating that the two distributions have become more dissimilar during fine-tuning.

Unexpectedly, running the same analysis with cosine similarities yields different outcomes. In the pretrained model, the PPI cosine similarities again have considerable overlap with the disease module (Figure S5a), though the disease module has a higher mean cosine similarity. Yet after fine-tuning, instead of seeing the PPI cosine similarities shift down, we observe in Figure S5a a positive shift in the PPI cosine similarities (where the mean increases from $0.253 \pm 0.001$ to $0.532 \pm 0.002$) as well as the disease module (where the mean increases from $0.313 \pm 0.001$ to $0.560 \pm 0.002$. We further see that the difference between the two distributions is no longer statistically significant (KS-test, $p = 0.18$).

If GF-Cardio is specific to cardiomyopathy, then we expect to see the AWs and the cosine similarities of the cardiomyopathy disease module to exhibit the most positive change in comparison to other diseases. In the attention weights, we do find that the AWs associated with the dilated cardiomyopathy disease module shifts up after fine-tuning, while most other disease modules shift downwards (Figure S5f). Indeed, out of a set of 163 diseases which have a disease module of at least 40 genes, only 2 (besides dilated cardiomyopathy) have higher median AWs after fine-tuning (Figure S5d). In contrast for cosine similarities, we observe that all disease module shift upwards after fine-tuning, not just the DCM disease module (Figure S5c,e). While the fine-tuned attention weights prioritize DCM above most other diseases, the fine-tuned embeddings do not show the same selective shifts.

### 6 Attention Weights Do Not Identify Physical Disease Gene Interactions

We next test whether Geneformer can capture the subtle distinction between a pair of disease genes, and an edge in the disease module. In other words, we ask if Geneformer can discern which interactions between disease genes are physical. To accomplish this we divide AWs into three categories: (1) AWs corresponding to all pairwise interactions between the 110 dilated cardiomyopathy (DCM) disease genes, whether or not they correspond to a physical interaction, (2) AWs corresponding to the 174 disease module edges, and (3) AWs corresponding to all physical interactions in the PPI. We aggregate these weights from both the pretrained and fine-tuned models. In the pretrained model we find that even though the model has not been trained to recognize cardiomyopathy, the attention weights within the disease module are higher than their counterparts in the PPI (Figure S5a). The disease module and disease genes have remarkably similar median values ($2.44 \times 10^{-3}$ and $2.63 \times 10^{-3}$ respectively) almost twice the median PPI AW ($1.43 \times 10^{-3}$). While both the disease module and the disease gene distributions are distinct from the PPI (KS test, $p = 2\times10^{-14}, 3\times10^{-231}$) they are statistically indistinguishable from each other (KS test, $p = 0.50$)

We see that even the pretrained core of Geneformer pays more attention to the disease module, which are important for determining the state of cells in which they are expressed. It does not however, offer a clear distinction between the physical interactions between disease genes (disease module edges, category 2) and the rest of the pairwise interactions between disease genes (category 1) (Figure S6a). We now ask whether fine-tuning the model can improve upon these distinctions.

While we see a clear prioritization of disease gene AWs during fine-tuning, we do not see any obvious separation between the fully connected, non-physical disease module, and the real disease module edges (Figure S6a). The two distributions are now statistically different according to the KS test ($p = 0.02$), but this distinction may not be a useful one.

We verify this by creating ranked lists of AWs, and using them to create a ROC curve. First, we combine the lists of disease module AWs and PPI AWs, and rank them in descending order. We then index down the list, and record the cumulative true positives (disease module edges). This naturally creates a ROC curve, where a perfect AUROC of 1 would be achieved if all the disease module edges comprised the highest attention weights, indicating a perfect separation of the disease module from the background (in this case the PPI). We repeat this process with the non-physical disease gene interactions as the background, and repeat the analysis with both the pretrained and fine-tuned models. We find that while fine-tuning improves our ability to distinguish the disease module from the PPI AWs (the AUROC goes from 0.70 to 0.81) it does not improve our ability to separate the disease module from the fully connected, non-physical disease module (AUROC goes from 0.53 to 0.55, see Figure S6b). In other words, while the fine-tuned model learns to prioritize genes involved in the cardiomyopathy disease module, it does not learn to prioritize the physical interactions between those genes.

The attention paid by the fine-tuned model to disease genes prompts us to ask how quickly attention to disease genes changes when we move away from the disease module in the network. In the cardiomyopathy disease module, we can find a path of at most 4 hops from any gene in the module to any other gene in the network. Most of these genes, even those only one or two hops away, will be effectively unrelated to the disease module [15]. We therefore ask what is the relationship between the attention weights of genes as a function of network distance from the disease module. To answer this question, we bin genes by their shortest path to the disease module (one hop, two hops, and so on). We then record the maximum attention between each of the disease module genes, and the genes in each bin. Given that there are 137 genes in the disease module, we will have 137 AWs per gene in each bin. We find that the interquartile range and median of the hop distance distributions declines as a function of increasing hop distance (Figure S6d), indicating that the magnitude of AWs captures the relative location of each gene in relation to the disease module. We find that a hop distance of 2 is comparable to randomly selected genes, replicating the average shortest path length between all genes in the PPI (ASPL = 2.6). Furthermore, we find that the hop distances below the ASPL have larger AWs than our random control while hop distances above the ASPL have smaller AWs.

#### 7 The Non-coding Interactome

We also tested Geneformer based disease module detection on an interactome that contains an additional 5,000 non-coding genes. This non-coding interactome (NCI) contains almost 500,000 additional regulatory interactions that are not present in the PPI, adding important biological context [3]. We find the same results in the NCI as the PPI, with an important caveat. After filtering for genes that are present in the 10,000 cell samples, we use to generate weight matrices, almost all of these non-coding interactions are removed, and the NCI becomes nearly identical to the PPI (Table S1). Where the PPI loses only 9% of its edges after filtering, the NCI loses 55% of its edges. Therefore, the networks become almost the same, causing the results to be trivially equivalent, although the absence of non-coding interactions from the 10,000 sample sets we analyzed merits further investigation.

| | PPI | | | NCI | | |
|---|---|---|---|---|---|---|
| | Nodes | Edges | $\langle k \rangle$ | Nodes | Edges | $\langle k \rangle$ |
| In GeneFormer vocab | 18,062 | 514,674 | 60 | 20,051 | 1,044,350 | 104 |
| In 10,000 sample set | 17,530 | 458,558 | 52 | 17,992 | 479,879 | 53 |

Table S1: Number of nodes and edges and average degree of the PPI and and NCI. Each network has more nodes and edges represented in the overall GeneFormer vocabulary than in a set of 10,000 randomly selected samples from Genecorpus, but the NCI loses far more edges than the PPI. After filtering for nodes and edges present in the samples, the networks are similar.

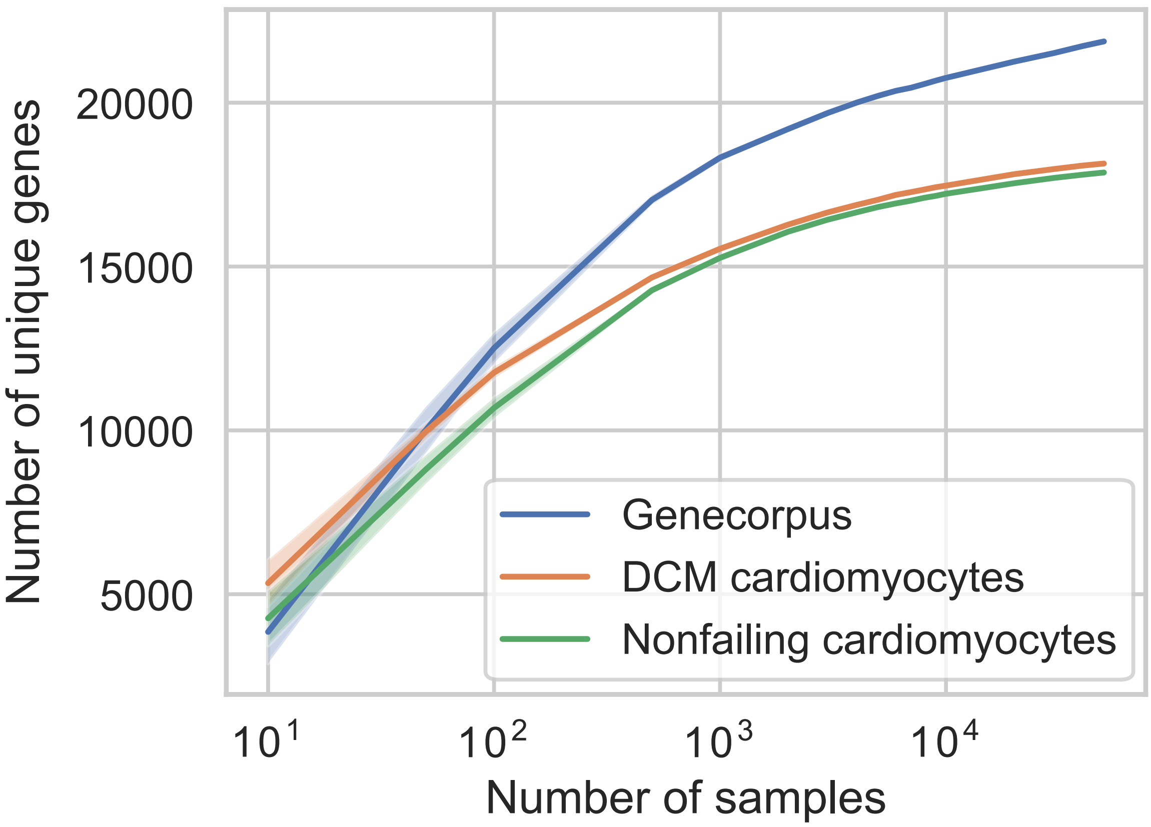

Figure 1: **Number of Unique Genes in a Sample Set** We find that the total number of unique genes in a set of $n$ samples scales sub-logarithmically with $n$. We select 10,000 samples because it is the largest number of samples we can feasibly use given computational constraints, and we confirm here that even adding thousands more samples would not substantially increase the number of genes we would find. Increasing to 50,000 samples, which would not be computationally feasible, would only increase the total number of observed genes by 5%. This is true whether we use Genecorpus, or cardiomycytes from the fine-tuned model's training data.

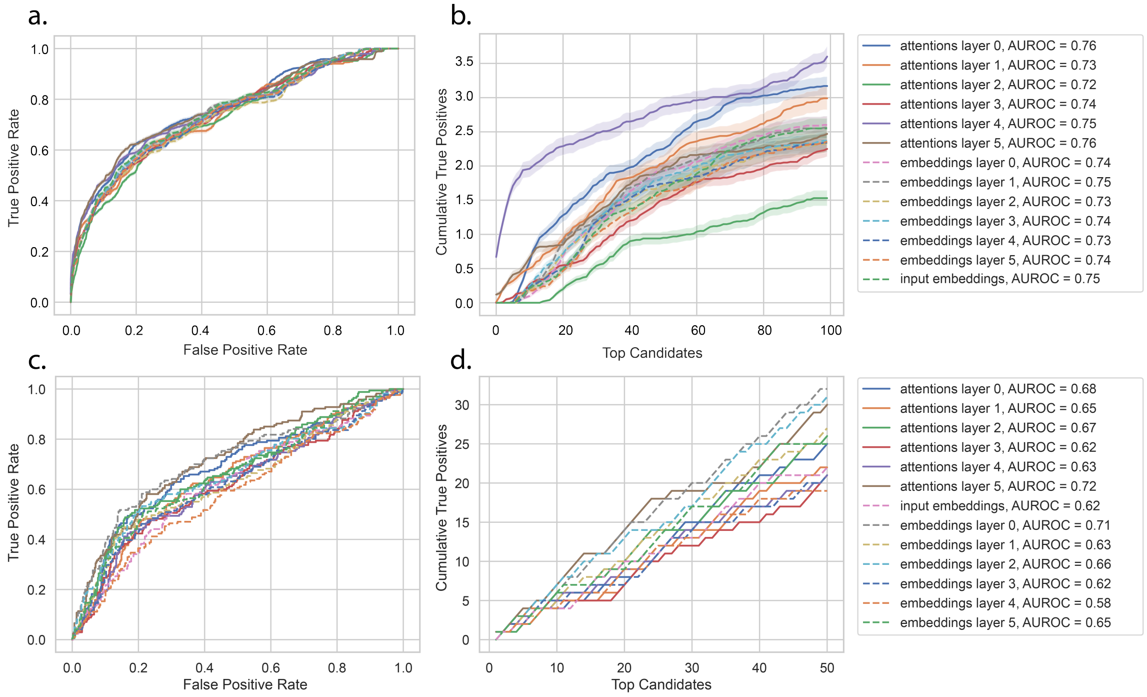

Figure 2: **Selecting a Layer in Geneformer (a)** ROC curves are plotted for disease module detection using networks weighted from each of Geneformer's 6 different layers (as well as the input embeddings). The highest AUROC is generated by the layer 4 attention weights. Ribbons represent the standard error in the mean over 100 realizations of the RWR. **(b)** The cumulative top hits in the top 100 candidates show that the layer 4 attention weights record the most hits in the top 50 candidates. **(c)** ROC curves are plotted for drug repurposing using networks weighted from each of Geneformer's 6 different layers (as well as the input embeddings). In this case the layer 5 attention weights generate the highest AUROC. **(d)** Cumulative top hits in the top 50 candidates are plotted for each network.

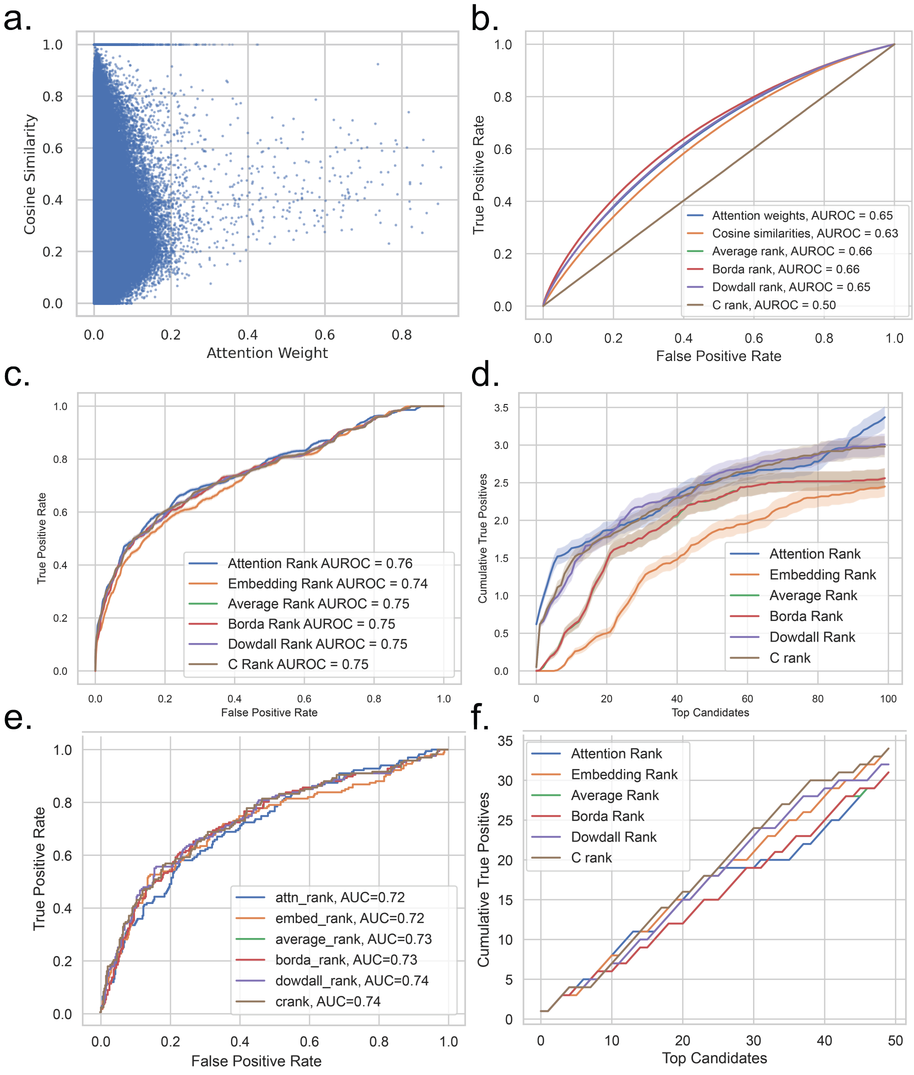

Figure 3: **Combining Attention Weights and Embeeddings (a)** Cosine similarities are plotted against embeddings for all pairs of genes in a 10,000 sample set. The pearson correlation between the cosine similarity between two genes and the attention weight is 0.16. **(b)** Attention weights and embeddings are ranked to separate PPI edges from background weights. Though both attention weights and embeddings perform better than random expectation, combining the rankings only yields 1% extra predictive power. **(c,d)** Using the best attention and embedding weighted networks for disease module detection (the layer 4 fine-tuned AW weighted network and the layer 4 fine-tuned embedding weighted network) we combine the rankings using several different methods. They do not provide additional predictive power. **(e,f)** The same procedure is implemented for drug repurposing. Here we combine the rankings from the layer 5 attention weighted network and the layer 0 cosine similarity network, and observe a 2% improvement in the AUROC, but no method records more true positives in the top 100 candidates than the layer 0 cosine similarity network.

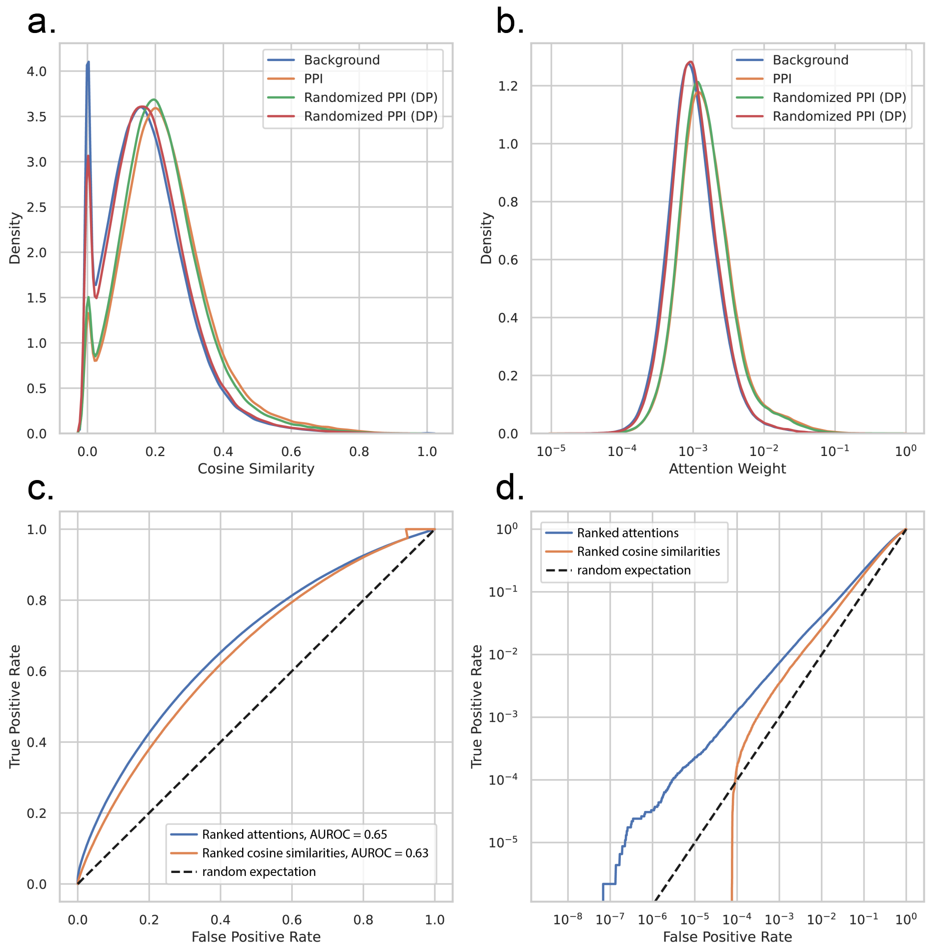

Figure 4: **PPI Edge Analysis (a)** Distributions of cosine similarities are plotted for the PPI, a degree preserving randomization of the PPI (PPI (DP)), a non-degree preserving randomization of the PPI (PPI (nDP)), and the background weights. **(b)** The same distributions are plotted for attention weights. **(c)** Using a list of ranked attention weights, ranked cosine similarities, and an averaged ranking, we plot how quickly we recover real PPI edges by indexing down the rankings. While all 3 methods perform better than random expectation, none recover more than 1% of the network in the first 10,000 weights. **(d)** The edge recovery ROC curves are plotted. The embeddings yield an AUROC of 0.63, the attentions 0.65, and the averaged rankings 0.66.

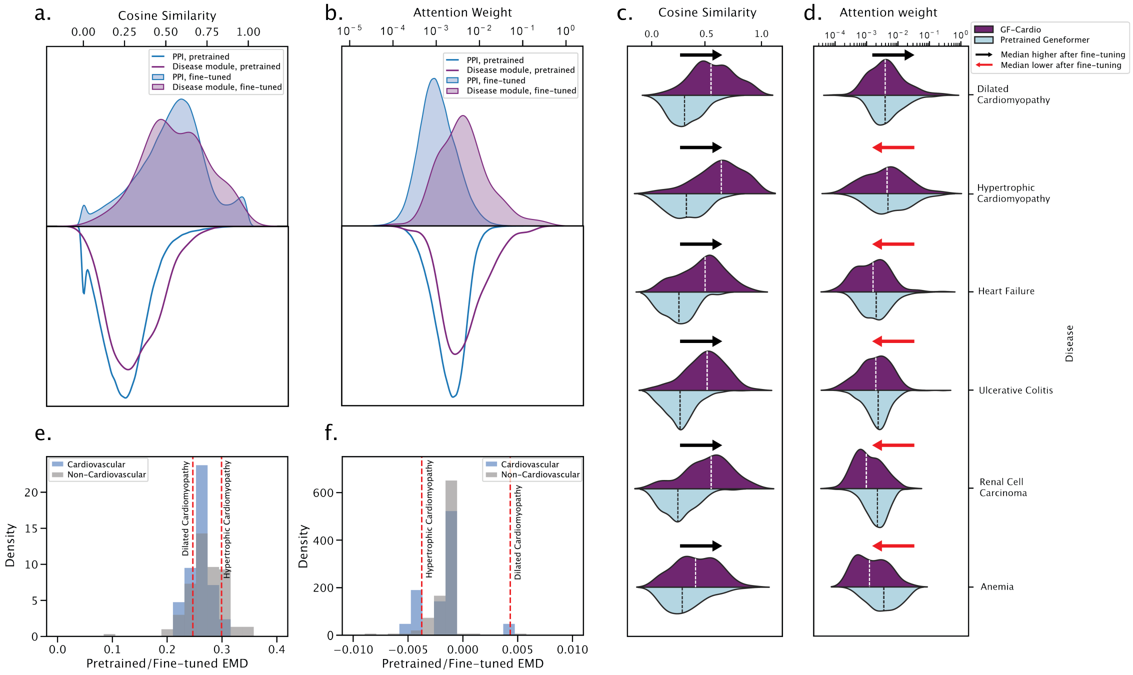

Figure 5: **Disease Modules in Geneformer (a)** Distributions of cosine similarities for PPI edges (blue) and disease module edges (purple) are plotted for the pretrained model and the cardiomyopathy model (shaded curves). **(b)** The same distributions are plotted for attention weights. **(c)** The histogram of earth mover's distances is plotted for 144 non cardiovascular diseases and 17 cardiovascular diseases. The EMD is calculated between the pretrained and fine-tuned distance of cosine similarities, and all diseases move up after fine-tuning. **(d)** The same histograms are plotted using attention weights. In this case, almost all disease modules move down after fine-tuning. **(e)** Distributions of cosine similarities for both models are plotted for several different disease modules. **(f)** Attention weights for both models are plotted for several different disease modules.

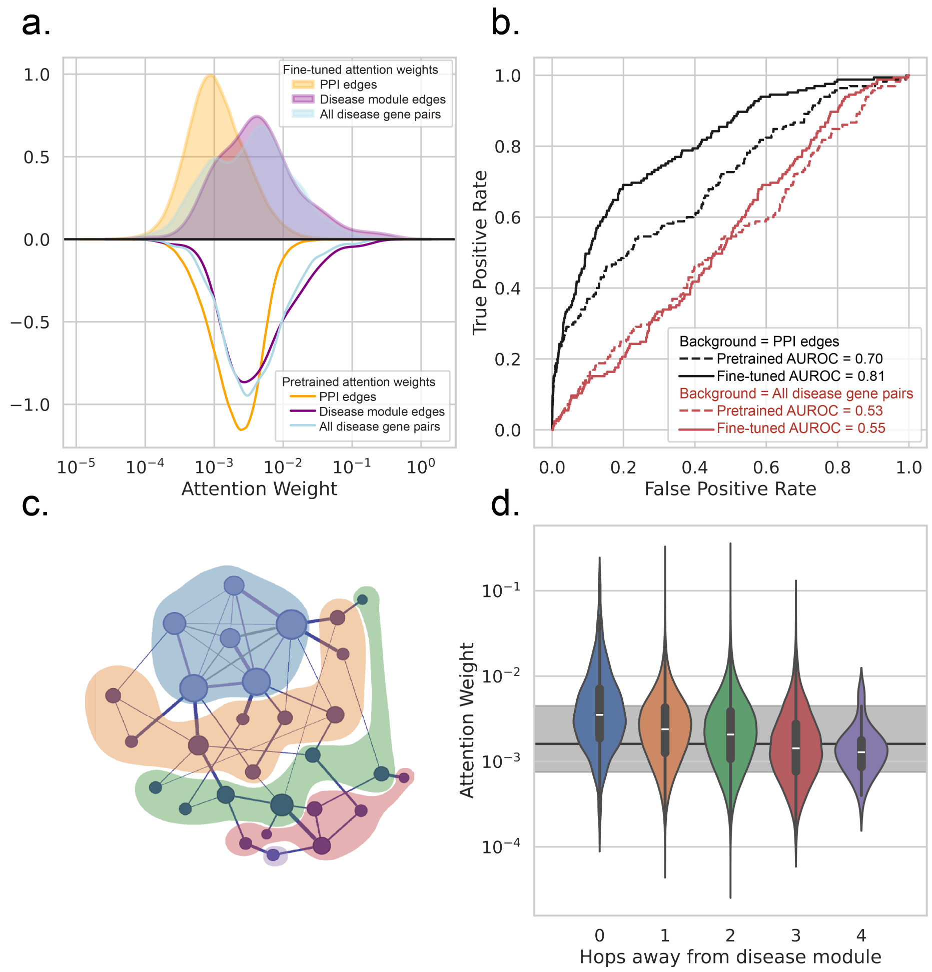

Figure 6: **Focusing on the Neighborhood of the Disease Module (a)** The distributions of attention weights are plotted for the pretrained and fine-tuned models. While the disease genes and disease module clearly separate from the PPI after fine-tuning, they do not differentiate from each other. **(b)** By using ranked lists of attentions from the pretrained model, we can separate the disease module from the PPI with an AUROC of 0.76, which increases to 0.82 after fine-tuning. However, we cannot tell apart the physical disease module interactions from non-physical interactions between disease genes either before (AUROC = 0.61) or after (AUROC = 0.6) fine-tuning. **(c)** The network neighborhood around the disease module (shaded in blue) is colored to show hop distance. The orange nodes are 1 hop away, the green nodes 2 hops, and so on. **(d** The attention on groups of genes that are n hops away from the cardiomyopathy disease module decreases as n increases. Above the average shortest path length in the PPI (2.6) the attention given to genes 3 and 4 hops away is lower than attention given to random genes.
